# Bright light therapy and early morning attention, mathematical performance, electroencephalography and brain connectivity in adolescents with morning sleepiness

**DOI:** 10.1101/2022.08.07.22278513

**Authors:** Martin H. Teicher, Elizabeth Bolger, Laura C. Hernandez Garcia, Poopak Hafezi, Leslie P. Weiser, Cynthia E. McGreenery, Alaptagin Khan, Kyoko Ohashi

## Abstract

Adolescents typically sleep too little and feel drowsy during morning classes. We assessed whether early morning use of an LED bright light device could increase alertness in school students. Twenty-six (8M/18F) healthy, unmedicated participants, ages 13-18 years, (mean 17.1±1.4) were recruited following screenings to exclude psychopathology. Baseline assessments were made of actigraph-assessed sleep, attention, math solving ability, electroencephalography and structural and functional MRI (N=10-11, pre-post). Participants nonrandomly received 3-4 weeks of bright light therapy (LiteBook Edge™) for 30 minutes each morning and used blue light blocking glasses for 2 hours before bedtime. LiteBooks™ were modified to surreptitiously record degree of use so that the hypothesis tested was whether there was a significant relationship between degree of use and outcome. LiteBooks™ were used 57±18% (range 23%–90%) of recommended time. There was a significant association between degree of use and: (1) increased beta spectral power in frontal EEG leads (primary measure); (2) greater post-test improvement in math performance and reduction in errors of omission on attention test; (3) earlier rise times on non-school days and reduced day-to-day variability in rise times; (4) increased dentate gyrus volume and (5) enhanced frontal connectivity with temporal, occipital and cerebellar regions during Go/No-Go task performance. Degree of bright light therapy use was associated with improvement in sleep cycle consistency, arousal, attention and functional connectivity, but not sleep onset or duration (primary measures). Although this was an open study, it suggests that use of bright morning light and blue light blocking glasses before bed may benefit adolescents experiencing daytime sleepiness.

## Introduction

As children pass through puberty, their circadian acrophase shifts, and they experience a strong urge to stay up and awaken late [1, 2]. High school typically starts early in the morning and a significant percentage of normal adolescents arrive at school each day with an insufficient amount of sleep [3-6], which can take a substantial toll on their academic performance [3, 7-10].

Relatively few studies have examined effects of sleep deprivation on cognitive performance in adolescents [11-15]. In these studies total sleep deprivation was associated with impaired memory performance and diminished computational speed [16, 17], while partial sleep deprivation was associated with deficits in reasoning [18], processing speed [15] and verbal creativity [19]. Some studies reported that simpler cognitive processes such as working memory and computational speed may not be significantly affected by a single night of sleep limited to 4 to 5 hours [19, 20]. However, even mild sleep restriction of an hour or more, when persistent across days, can lead to memory problems as severe as seen following total sleep deprivation [21]. In general, cumulative sleep deficits are associated with impaired verbal memory consolidation [22], deficit in vigilance, processing speed and working memory [23] and poor performance on serial digit-learning tests during morning but not afternoon test sessions [24].

Between 58-68% of high school students surveyed in Ontario report that they feel “really sleepy” between 8 and 10 A.M. [7]. Thus, achievement in early morning classes may suffer the most in sleep-deficient adolescents.

Fortunately, sleep only needs to be extended by a modest amount to enhance cognition in children. Sadeh et al. [25] showed that performance on memory, attention and vigilance tasks improved significantly after 1 hour of sleep extension on three consecutive nights. Gais et al [26] and Backhaus et al [27] have also shown the beneficial effects of sleep on memory consolidation in children and adolescents.

As the primary reason for insufficient sleep is a naturally occurring propensity to stay up later in the evening [1, 2, 28] it seems plausible that bright light treatment (BLT) at the appropriate time may phase advance biological clocks and potentially reverse this problem [29, 30]. Hence, we sought to test the hypothesis that consistent morning use of an LED BLT device (LiteBook Edge™) by healthy adolescents would shift the phase of their sleep wake cycle and enable them to receive an increased amount of sleep during the school week and perform better on tests of attention and academic performance and evidence signs of improved alertness.

Alternatively, BLT could potentially enhance alertness through other mechanisms, such as a direct arousing effect, without exerting a discernible effect on circadian phase or sleep duration [31, 32].

## Methods

### Participants

All but two participants were enrolled in the Walnut Hill School, where a call for participants and an explanation of the study was made during a general assembly. The other participants were children of Walnut Hill staff. A specific school was selected to enable on-site neurocognitive and electroencephalographic assessments of individuals right before they began morning classes. Participants were normal adolescents of either sex, age 13 -18 years who were unmedicated and showed no symptoms of psychopathology or circadian rhythm sleep disorder. This study was reviewed and approved by Partners Health Care IRB. Parents or guardians of participants under age 18, who were interested in participation, provided written informed consent and youths provided written assent. Eighteen-year-old participants provided written informed consent. Subjects were required to have an IQ greater than 80 [33], and to be enrolled in a normal academic program. Individuals who participated in athletic training or performance art prior to the start of the normal school day or engaged in other activities that could markedly influence morning alertness were ineligible. Participants needed to indicate that in their opinion their academic potential exceeded their current performance, that they experience some degree of morning sleepiness and that they would be willing to consistently utilize a device that might enable them to perform better at school. All individuals were paid for their participation. Our intent was to recruit thirty-six participants based on power analysis, with N=20 choosing to participate in the neuroimaging subsample.

### Design

This was a nonrandomized study in which all participants received the BLT device. However, the device was modified to surreptitiously record degree of use so that the hypothesis that we sought to test was whether there was a significant dose-dependent relationship between degree of BLT use and outcome. The study was conducted between March 27, 2017, and June 23, 2017. The protocol ID is 2016D003724 and the clinicaltrials.gov ID is NCT05383690. The protocol was registered after the trial took place as at the time of approval in 2016 only trials funded by NIH were eligible for registration on clinictrials.gov. The authors confirm that all ongoing and related trials for this intervention are registered

Baseline assessments included seven days of activity monitoring (ActiGraph Link GT9X) to determine bedtime, rise time and total sleep time during the school week and weekends using the Cole-Kripke algorithm [34]. Testing took place at the end of the school week (Thursday, Friday or Saturday AM) to assess the cumulative effects of sleep restriction. They were tested on the Quotient ADHD System [35] to assess attention. The Permanent Product Measure of Performance (PERMP) test was used as a treatment-responsive measure of academic performance and processing speed in mathematics [36], along with a sleep-deprivation sensitive Serial Addition/Subtraction Task (SAST) [37]. These tests were selected as easily captured and quantified measures of attention, computational speed and higher-level mathematical ability given the concern that morning drowsiness may have on academic performance during morning classes.

During the 3^rd^-4th weeks of BLT, participants rewore actigraphs and were evaluated at the end of the school week using Quotient, PERMP, and SAST.

### Behavioral and Emotional Screening System from the BASC-2

The Self-Report of Personality component of the BASC-2 was used for screening. It includes clinical scales for anxiety, atypicality, locus of control, social stress, somatization, attitude toward school, attitude toward teachers, sensation seeking, depression, and sense of inadequacy. For inclusion we required participants to be unmedicated and score within the normal range on the BASC-2.

### Modified Walter Reed serial addition/ subtraction task (SAST)

Participants were presented, by computer, with two single-digit numbers in succession, followed by an operator (“+” or “–”). Each item was presented for 200 ms with a 200 ms delay between items [38]. Participants selected their response on a display slider. Each session consisted of 50 trials. Data analyzed were correct response latencies.

### Permanent product measure of performance (PERMP)

A collection of high school placement exam questions in mathematics was used to measures participant’s ability to pay attention, stay on task and perform calculations up to their level of ability. Improvement was demonstrated by an increase in the number of attempted and successfully completed problems. Children were given 10 minutes to complete as many problems as they could in the allotted time. There were two equivalent forms of the test, and each contained 62 problems. Children took one form as a pre-test and the other as post-test.

### Quotient ADHD system (Quotient)

This was a commercially available evaluation platform developed by the PI [35, 39] and marketed at the time by BioBehavioral Diagnostic Systems, a division of Pearson. Briefly, the system consisted of an infrared motion analysis system that tracked the participant’s head movements while they performed a monotonous but demanding cognitive control task. During this task subjects were instructed to respond as rapidly and as accurately as possible by key press to 5-, 8- and 16-pointed stars that appeared at random screen positions and to not respond to 4-pointed stars. Approximately 90% of the stimuli were targets. Stimuli were presented for 240 msec with a variable interstimulus interval (ISI) (mean 2500 msec) [40]. Standard response measures consisted of accuracy, errors of commission, errors of omission, response latency and variability in correct response latency.

### Epworth sleepiness scale (ESS)

The ESS is a self-administered questionnaire in which respondents indicate their perceived likelihood of dozing off or falling asleep while engaged in eight different activities [41, 42]. Internal consistency (Cronbach’s alpha = .88) and test-retest reliability (.82) are good [41]. ESS scores correlate with multiple sleep latency test scores and vary in the expected manner between healthy controls and individuals with narcolepsy, obstructive sleep apnea and idiopathic hypersomnia [42]. It is important to note that the ESS focuses on an individual’s potential to doze off and not their feelings of fatigue or drowsiness/sleepiness, which are different but related concepts [43-45].

### Electroencephalography (EEG)

Traditionally, EEG has served as the gold-standard for assessing alertness, drowsiness and sleep, and EEG spectral parameters can provide a minute-to-minute index of vigilance and alertness [46, 47]. In general, drowsiness or decreased alertness is characterized by enhanced slow wave activity, particularly theta [48-50], and decreased higher frequency activity, particularly beta [51-53]. EEGs were recorded on a HP ProBook 4730s (IntelCorei7-2630QM) running WinEEG Version 2.96.63 (03.2014) (Mitsar Co. Ltd, 197374 St. Petersburg, Russia) using a 25-channel Mitsar-EEG-201 amplifier with standard 19-channel Electro-Gel caps (Eaton, Ohio 45320) with impedance < 5 KΩ and Monopolar1 (A1<->A2) montage following manufacturer recommendations.

Acquisitions consisted of 3 minutes eyes closed and 3 minutes eyes open EEG at with a 2 min break between sessions. Portable EEG equipment was brought to the school so that EEG could be collected prior to their first morning class. The two participants who did not attend the Wallnut Hill School were monitored at the laboratory. All EEG collections took place between 7 AM – 9 AM on Thursdays, Fridays or Saturdays during baseline week and were repeated during the final week of BLT. Participants did not receive BLT prior to their post-treatment EEG and evaluation.

Color images were created in R using generalized additive models (GAM) with integrated spherical spline smoothness estimations and plotted in ggplot2 using methods described online (https://stackoverflow.com/questions/35019382/topoplot-in-ggplot2-2d-visualisation-of-e-g-eeg-data/35026677).

### MRI acquisition

Participation in the MRI portion of the study was optional. At the end of this baseline week interested participants were scanned between 7 – 9 AM with sequences for morphometry, resting state and task-based functional connectivity. Multi-band sequences were used to reduce acquisition time to better control for physiological confounds by provide scanning rates higher than respiratory rates.

During the 10-minute task-based scan they performed a version of the Quotient ADHD system vigilance task. Scanning took place prior to initiation of BLT and again at the end of the treatment period, but BLT was not used on the day of scanning.

Scans were performed on either a Siemens 3T TIMS Trio MRI system using a 32-element head RF coil, or on a Siemens 3T Prisma MRI system with a 64-element head RF coil using the same sequences. Performance and stability were equivalent on these systems. Two different scanners were used to ensure that scanning took place early in the morning and at the same times for their pretreatment and posttreatment scans. Subjects were retested on the same machine at the same time at the end of the protocol.

The resting-state scan parameters were: timing: TE/ TR / images = 30ms / 0.750sec / 800 volumes; Resolution: voxel size / matrix = 2.75×2.75×2.8 mm^3^ voxels, FOV = 80×80×54. Acquisition: GRAPPA = 2, multi-band acceleration = 6, echo spacing = 0.51 ms. flip angle = 52 deg.

Both T1 and T2 contrast images were acquired in 3D for volumetric analysis. Parameters for the multi-echo T1 contrast “magnetization prepared rapid gradient echo” (MPRAGE) sequence were TE (4) /TR / TI / duration = 1.7, 3.5, 5.4, 7.3ms / 2.5sec / 1.2 s / 4:42. Resolution: voxel size / matrix = 1.0×1.0×1.33 mm^3^, FOV = 256×256×170. Acquisition: GRAPPA = 2, echo spacing = 10ms, flip angle = 7deg. Parameter for the fast spin echo T2 contrast “sampling perfection with application-optimized contrasts by using flip angle evolution” (SPACE) were: TE / TR / duration = 308ms / 2.8sec / 2:14 (Trio) or 1:32 (Prisma). Resolution: voxel size / matrix = 1.0 × 1.0 × 1.33 mm^3^, FOV = 256×192×170mm. Acquisition: GRAPPA = 2, echo spacing = 3.2ms, flip angle = variable.

### MRI analysis

Volumetric segmentation was performed with the Freesurfer image analysis suite using both MPRAGE and T2 SPACE sequences. Brain regions were segmented and labeled using the ‘recon-all’ pipeline, which included motion correction, non-brain tissue removal [54], Talairach transformation, deep gray matter and subcortical white matter volumetric segmentation [55, 56], intensity normalization [57], tessellation of white matter / gray matter boundaries, topology correction [58, 59], and surface deformation following intensity gradients to optimally locate gray/CSF and gray/white borders [60-62]. We focused entirely on the dentate gyrus. Hippocampus subfields were extracted using segmentation procedures included in version 6 [63]. Overall, this approach provided hippocampal subfield volume measures that aligned more closely with histological measurements than with the prior FreeSurfer release or alternative automated segmentation algorithms [63]. The aim was to determine if consistent use of morning LED BLT was associated with increase in dentate gyrus volume, as studies in rats have shown that 4 weeks of 10K phototherapy (30 min/day) increased dentate gyrus neurogenesis [64]. Seed-based functional connectivity analyses were performed using the CONN toolbox [65]. Preprocessing included estimation and correction for head motion, slice timing correction, temporal and spatial normalization in Montreal Neurological Institute space, and smoothing using an isotropic 8 mm half-maximal Gaussian kernel. The CompCor method [66] was used to address spike and motion artifacts. White matter and cerebrospinal fluid principal components and realignment parameters were entered as confounds in a first-level analysis, and data were band-pass filtered to 0.008 to 0.09 Hz. This method addresses the confounding effects of participant movement, without regressing the global signal and without affecting intrinsic functional connectivity [67]. Given the limited sample size we sought to assess was whether there were significant alterations in prefrontal functional connectivity that correlated with degree of BLT use. We hypothesized that BLT enhancement in connectivity would be more apparent during the attention task as it would likely bring prefrontal interconnections into play to facilitate performance. Neuroimaging data from this study has been uploaded to OpenNeuro. doi:10.18112/openneuro.ds004219.v1.0.0

### LED bright light therapy (BLT) device

Subjects were provided with the LiteBook Edge™ (LiteBook Company LTD) for individual use at home. This is a patented smart phone sized BLT device that provides 10,000 lux illumination at a recommended distance of 61 cm from an LED panel with peak spectral radiance in the blue color spectrum that closely corresponds to the peak spectral frequency (480 nm) of melanopsin photoreceptors that project to the suprachiasmatic nucleus and entrain the circadian clock [68]. Subjects were instructed to use the BLT device, as early as possible, for 30 minutes each morning during the 4 week trial. These devices were equipped with monitoring electronics that enabled us to download their daily degree of use when they completed the study. Participants were also provided with yellow-tinted blue light blocking glasses and were instructed to wear them starting 2 hours before bedtime if they were viewing LED or LCD screens. Subjects were also provided with their own LiteBook Edge™ and blue blocking glasses to keep at the end of the study.

### Statistical analysis

Data were analyzed using linear mixed effect or ANCOVA models with two-tailed significance levels to test hypotheses that greater use of BLT would be associated with phase advance, increased sleep time, improved measures of vigilance and mathematical performance, increased beta and gamma EEG activity, decreased theta EEG activity, increased dentate gyrus volume, and increased prefrontal functional connections during Go/No-Go task performance. The unit of analysis were individual participants pre- and post-treatment measures. Before starting the analyses, we selected beta and theta EEG activity, and sleep onset and sleep duration as primary measures. Errors of omission and response variability on the Quotient ADHD System, mathematical ability on the PERMP, computational speed on the serial addition/subtraction task, dentate gyrus volume, and sleep propensity on the Epworth Sleepiness Scale were considered secondary measures. Changes in frontal pole functional connectivity were considered exploratory outcomes given the small sample size. Research staff collecting and processing data were blind to percent use data until the final analyses were run. Statistical analyses were conducted in R version 4.0.3. Scripts for all the reported analyses, output tables and data files are included in the supplementary materials.

Measures collected were identical to what we proposed in our detailed IRB protocol, which is included in the supplementary materials. Statistical analyses closely followed what we originally proposed in the IRB protocol except that we eliminated some of the proposed analyses given the smaller than anticipated sample size. We eliminated analysis of mean activity during the first 2 hours after awakening, percent time spent fully attentive, maximum fractal length of the EEG waveform and global field power of upper alpha band (10-12 Hz) oscillations. We substituted more traditional measures of alpha and gamma spectral activity. We also proposed to make 42 functional connectivity comparisons but limited the functional connectivity analyses to two regions, the right and left frontal poles based on the EEG findings.

## Results

### Participants

As illustrated in the CONSORT flow chart (Fig 1) twenty-six individuals (8M/18F, ages 13.1-18.8 years, mean 17.1 ± 1.4 years) were screened and all were enrolled in the study. Seventy-four percent of the subjects were White, 11% Asian, 7% Black and 1 participant was a Native American. Average composite IQ on the K-BITS-2 was 111 ± 15, with mean verbal score of 110 ± 14 and non-verbal score of 108 ± 15. Based on the ESS, 19%, 35%, 23%, 19% and 4% of participants had low normal, high normal, mild excessive, moderate excessive and severe excessive daytime sleep propensity, respectively.

**Fig 1.**
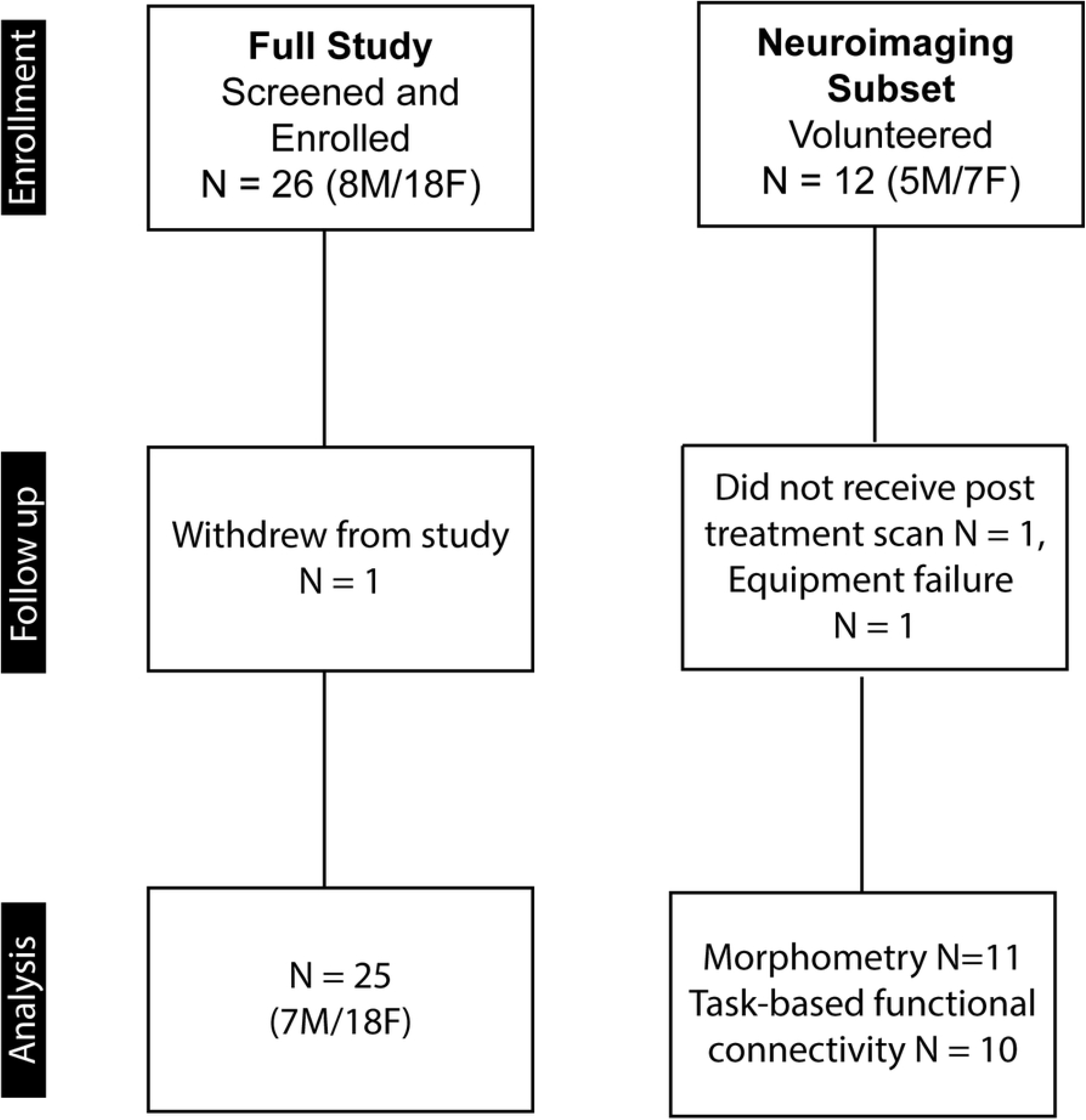
CONSORT flow diagram. Sample sizes for the enrollment, follow-up and analysis phases of the full study and the neuroimaging subset.

### LED phototherapy utilization

One participant did not use the device and withdrew from the study after the initial assessment. This participant was excluded from the analyses, as carrying his initial ratings forward, coupled with 0% device use, would have inflated the significance of the findings. On average the remaining participants used the device 57 ± 18 percent of the time; ranging from 23% to 90% of recommended use. Data analyses were conducted on N=25 pre / post measures.

### Sleep parameters

Dates were categorized into school and non-school days. Nap periods constituted 8.4% of detected sleep periods and were excluded from the analyses. Three of the participants had bedtimes that were consistently after midnight, but all participants were able to rise at desired times on school days. Hence, none met criteria for Circadian Rhythm Sleep-Wake Disorders - Delayed sleep phase type.

There was a significant interactive effect between percent of BLT use and school day category on rise time (F_1,308_ = 6.76, p= .0098), with greater use of the device leading, in particular, to earlier rise times on non-school days. Further, device use was associated with reduced day-to-day variability in rise times (Likelihood ratio (LR) test = 18.51, p < .0002).

Contrary to our expectation there was no significant effect of percent device use on bedtime (F_1,308_ = 1.31, p = .25) or sleep onset time (F_1,310_ = 1.42, p = .23). Total sleep time was influenced by age (F_1,24_ = 6.32, p = .019) but there was no significant effect of percent BLT device use (F_1,310_ = .60, p = .44), though there was a trend for total sleep time to be more consistent with BLT (LR Test = 4.68, p = .06). There were also trend level association between degree of device use and enhanced sleep efficiency (F_1,307_ = 3.39, p = .066) and reduction in ESS scores (F_1, 23_ = 3.81, p = .063). There was a significant reduction in ESS scores with BLT in the 12 participants with increased levels of sleep propensity at baseline (t_10_= 2.95, p = .015).

### Attention and cognitive performance

Performance on the SAST was strongly associated with the difficulty of the calculation, but there was no significant effect of percent BLT device use on SAST accuracy (F_1,2411_ = 0.342, p = .56). However, BLT use was associated with a 22% reduction in response latency variability (LR Test = 63.61, p <.0001), which is indicative of enhanced vigilance. Percent of device use was positively associated with number of math questions answered correctly on the PERMP (F_1,22_ = 5.25, p = .031), which was not influenced by differences in baseline performance (LR test = .82). The association between percent device use and percent change in PERMP scores was large, accounting for 19.2% of the variance in percent change scores (90% CI 0.01-0.42).

As seen in Table 1 greater use of BLT was associated with increased post-test improvement in accuracy and reduction in errors of omission on the Quotient ADHD System with medium and large effect sizes, respectively. Interestingly, percent BLT use was associated with a reduction in time spent immobile percentile score (beta = -0.387, p = .035), which is consistent with a reduction in drowsiness.

**Table 1.**
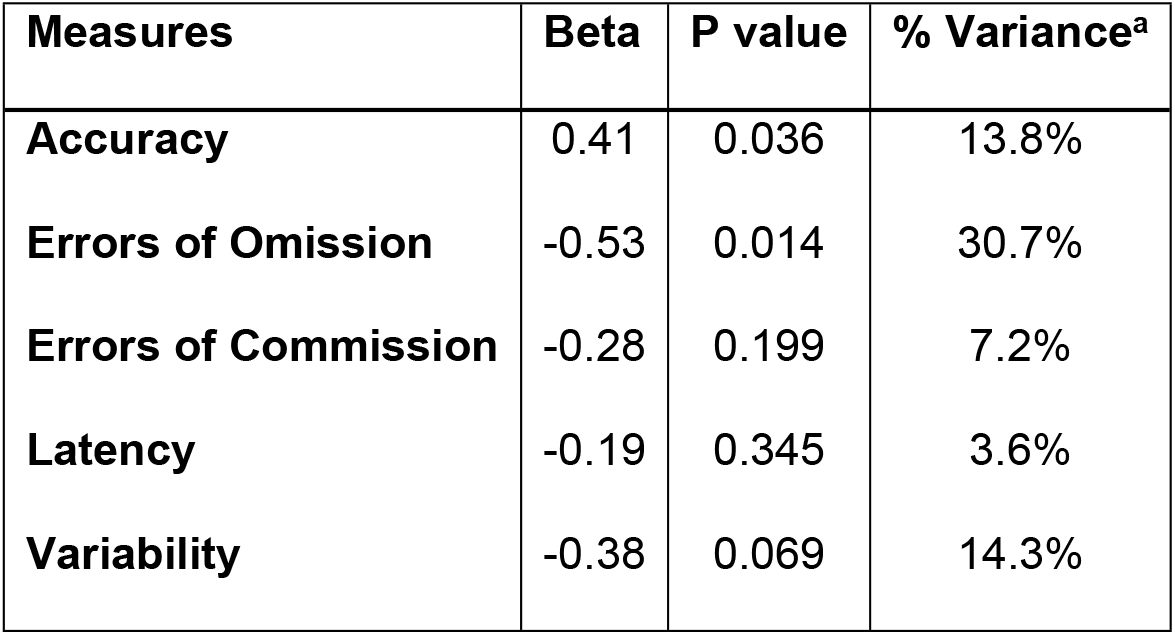
Bright light therapy use and Quotient attention measures.

Association between percent LED phototherapy device use on pre-treatment – post-treatment measures of attention and vigilance on the Quotient ADHD system. ^a^Effect size (percent variance accounted for) was determined using the variance decomposition method of Lindeman, Merenda and Gold [69, 70].

### Electroencephalography

Fully useable pre-post eyes open EEGs were available on 20 participants. After artifact rejection, EEGs were analyzed to extract mean absolute spectral power in delta (1.5 – 4 Hz), theta (4 – 7.5 Hz), alpha (7.5 – 14 Hz), beta1 (14 – 20 Hz), beta2 (20 – 30 Hz) and gamma (30 – 40 Hz) frequency bands. As seen in the statistical maps in Fig 1, degree of device use was significantly associated with increase in eyes-opened beta1 spectral power in leads Fp1 (F_1,18_ = 9.38, p = .0067, 16.5% of variance) and Fp2 (F_1,18_ = 19.20, p = .0004, 25.5% of variance), and also (not shown) in beta2 spectral power in the same leads (F_1,18_ = 5.34, Fp1: p = .033, 7.6%; Fp2: F_1,18_ = 6.72, p = .018, 12.9%), as well as alpha in Fp2 (F_1,18_ = 8.20, p = .01, 14.4%) and gamma in Fp2 (F_1,18_ = 7.03, p = .016, 7.6%) and C3 (F_1,18_ = 5.69, p = .018, 9.1%).

Least square estimated influence of device use on EEG spectral topography from the mixed effects models is illustrated in Fig 2, which shows estimated spectral power prior to the initiation of BLT and then following 50% and 80% of prescribed use. Curiously, there were no significant reductions in delta or theta activity that correlated with degree of device use, nor were there significant increases in beta/theta ratio.

**Fig 2.**
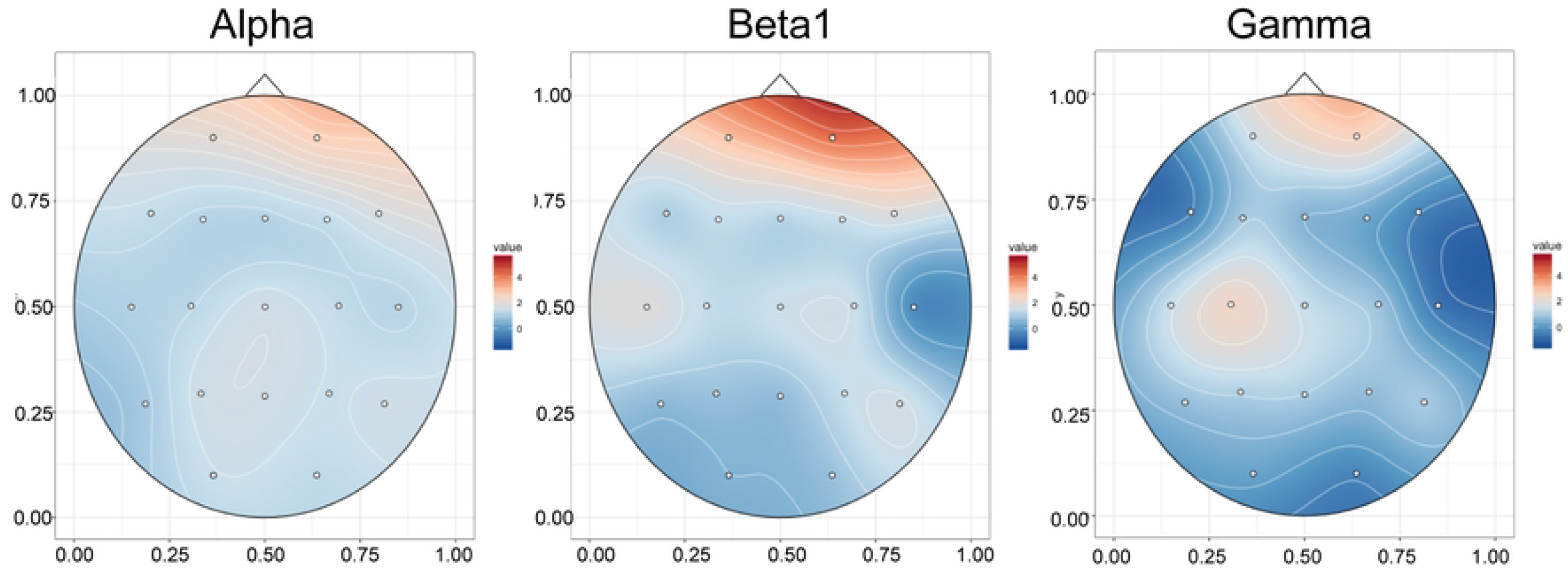
Association between bright light therapy use and eyes open EEG spectral activity. Topoplots of T-scores showing statistical association between degree of use of an LED bright light therapy device and percent increase in eyes-open Alpha (7.5–14 Hz), Beta1 (14–20 Hz) and Gamma (30–40 Hz) spectral activity.

There was less apparent association between degree of BLT use and eyes-closed EEG spectral power in the 22 participants with fully useable data. As seen in Fig 3, increase in Beta1 (F_1,20_ = 8.29, p = .0093, 5.6% variance explained) and Beta2 spectral activity (F_1,20_ = 8.5, p = .0085, 9.3% variance) in Fz were significantly associated with degree of device use.

**Fig 3.**
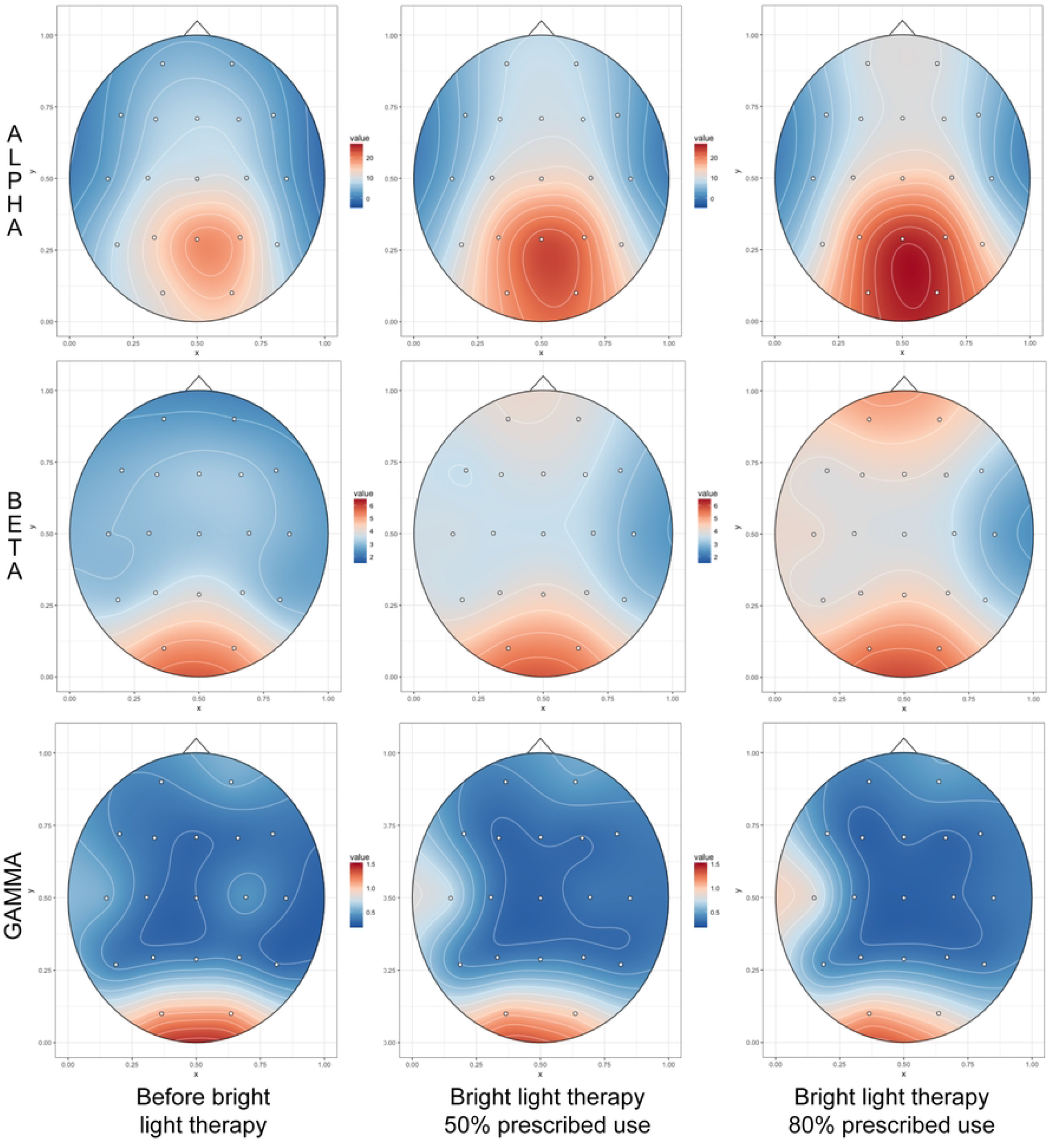
Distribution of eyes open EEG spectral activity based on percent use of bright light therapy. Topoplots of the distribution of Alpha, Beta1 and Gamma spectral activity prior to initiation of LED BLT device use and at 50% and 80% of prescribed use based on linear mixed effects regression models.

### Functional Connectivity

Overall, twelve subjects participated in the MRI component of this study, but only 11 (4m/7F) completed both scans and the display projector failed during one of the post-test scans on a female participant, so exploratory results are based on N=10 pre/post fMRI scans. To assess whether percent BLT device use enhanced visual attention we focused on the connectivity of the left and right frontal poles as EEG indicated that the largest changes in spectral activity were associated with these regions. As seen in Table 2 and Figs 5 and 6, degree of device use was associated with increased connectivity between the left frontal pole and two clusters. One contained the posterior cingulate and the precuneus. The other contained the cerebellum crus 2. Degree of device use was also associated with enhanced connectivity of right frontal pole with five clusters. The first was a large cluster that contained the cerebellum crus1, cerebellum 6, fusiform gyrus, lingual gyrus, intracalcarine cortex and occipital pole. Connectivity was also increased to the anterior and posterior portions of the medial temporal lobe and the inferior frontal gyrus operculum of the left hemisphere. Finally, there was a graded increase in connectivity between the right frontal pole seed and other portions of the right and left frontal pole.

**Table 2.** Changes in Functional Connectivity with percent use of bright light therapy.

| Regions | Hemisphere | x | y | z | Size | Size pFDR<sup>a</sup> | Peak p- |
| --- | --- | --- | --- | --- | --- | --- | --- |
| ***Left Frontal Pole Seed*** |  |  |  |  |  |  |  |
| Cerebellum crus 2 | Left | -24 | -90 | -34 | 244 | 0.000325 | 0.000002 |
| Posterior cingulate, precuneus | Left | +18 | -44 | +38 | 144 | 0.006866 | 0.000010 |
| ***Right Frontal Pole Seed*** |  |  |  |  |  |  |  |
| Cerebellum crus 1, 6, lingula gyrus, occipital pole, fusiform gyrus, intercalcarine cortex | Left | -10 | -80 | -24 | 587 | 0.000000 | 0.000016 |
| Frontal pole | Right, Left | +06 | +66 | +28 | 248 | 0.000060 | 0.000007 |
| Inferior frontal gyrus operculum | Left | -48 | +12 | +08 | 106 | 0.013547 | 0.000048 |
| Middle temporal gyrus, posterior | Left | -60 | -50 | +02 | 96 | 0.013916 | 0.000006 |
| Middle temporal gyrus, anterior | Left | -50 | -22 | -12 | 95 | 0.013916 | 0.000028 |

Pretreatment – post-treatment changes in connectivity of left and right frontal poles during performance of a go / no-go cognitive task that correlated with degree of BLT device use.
<sup>a</sup>FDR – False Discovery Rate statistical correction for cluster size.

**Fig 4.**
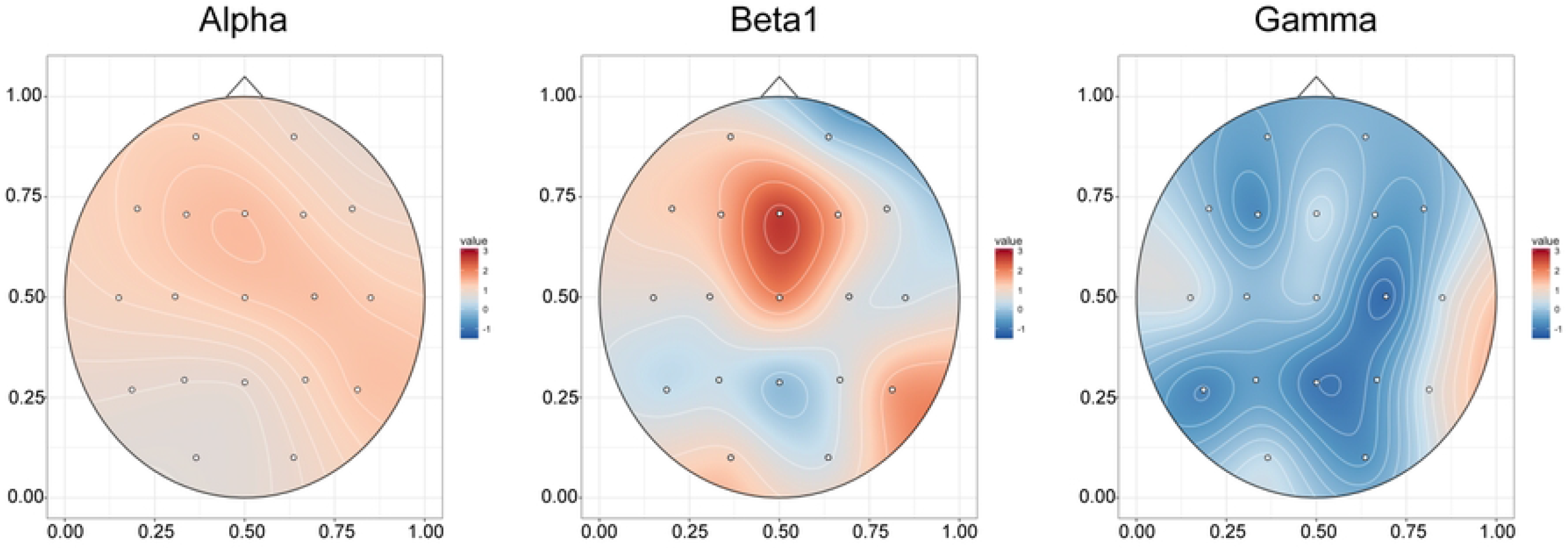
Association between bright light therapy use and eyes closed EEG spectral activity. Topoplot of T-scores showing statistical association between degree of morning BLT device use and percent increase in eyes closed absolute Alpha (7.5–14 Hz), Beta1 (14–20 Hz) and Gamma (30–40 Hz) spectral activity.

**Fig 5.**
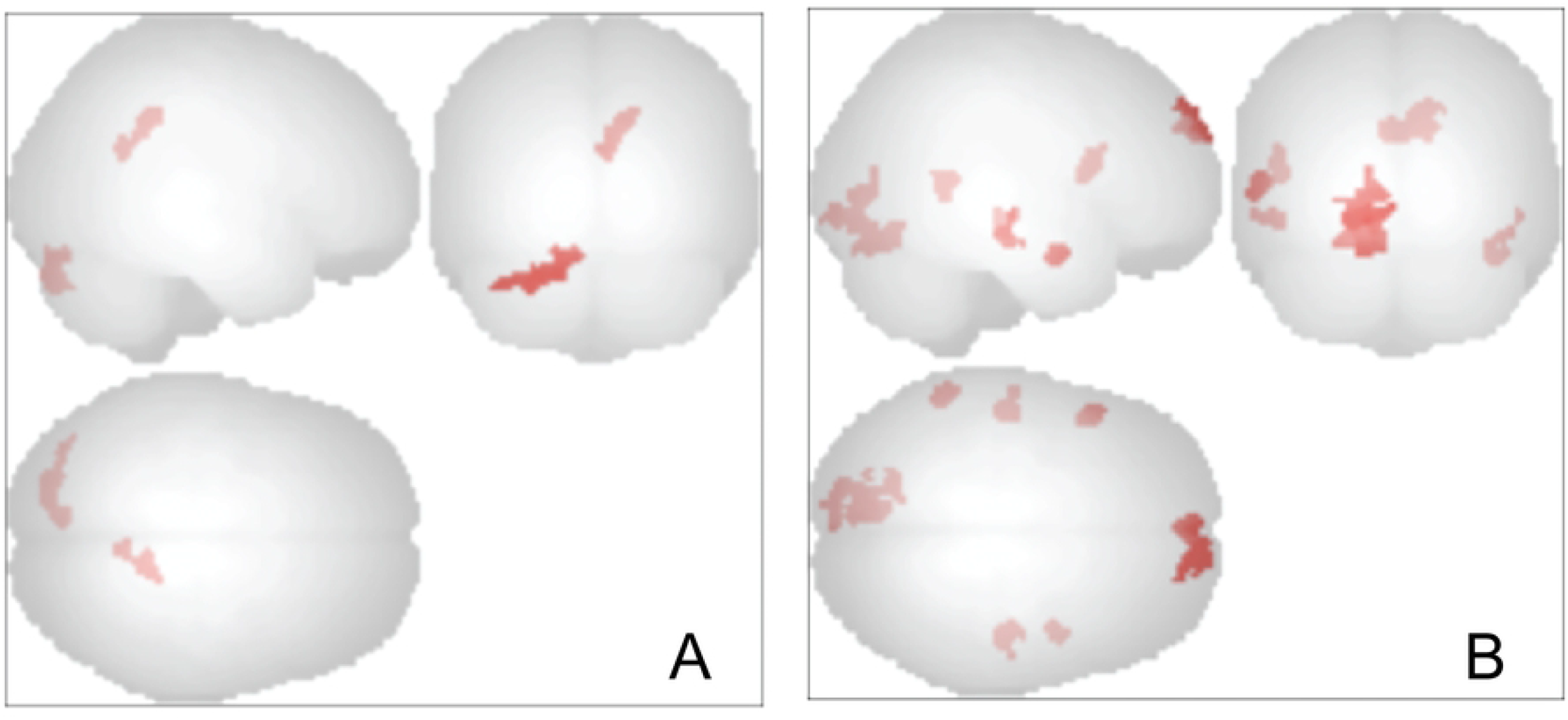
Change in functional connectivity of frontal poles in relation to degree of use of bright light therapy. **A**. Clusters with increased functional connectivity to the left frontal pole that correlated with degree of bright light therapy use. **B**. Clusters with increased functional connectivity to the right frontal pole that correlated with degree of bright light therapy use. Height threshold p < .001 uncorrected, cluster size p < .05 FDR corrected.

**Fig 6.**
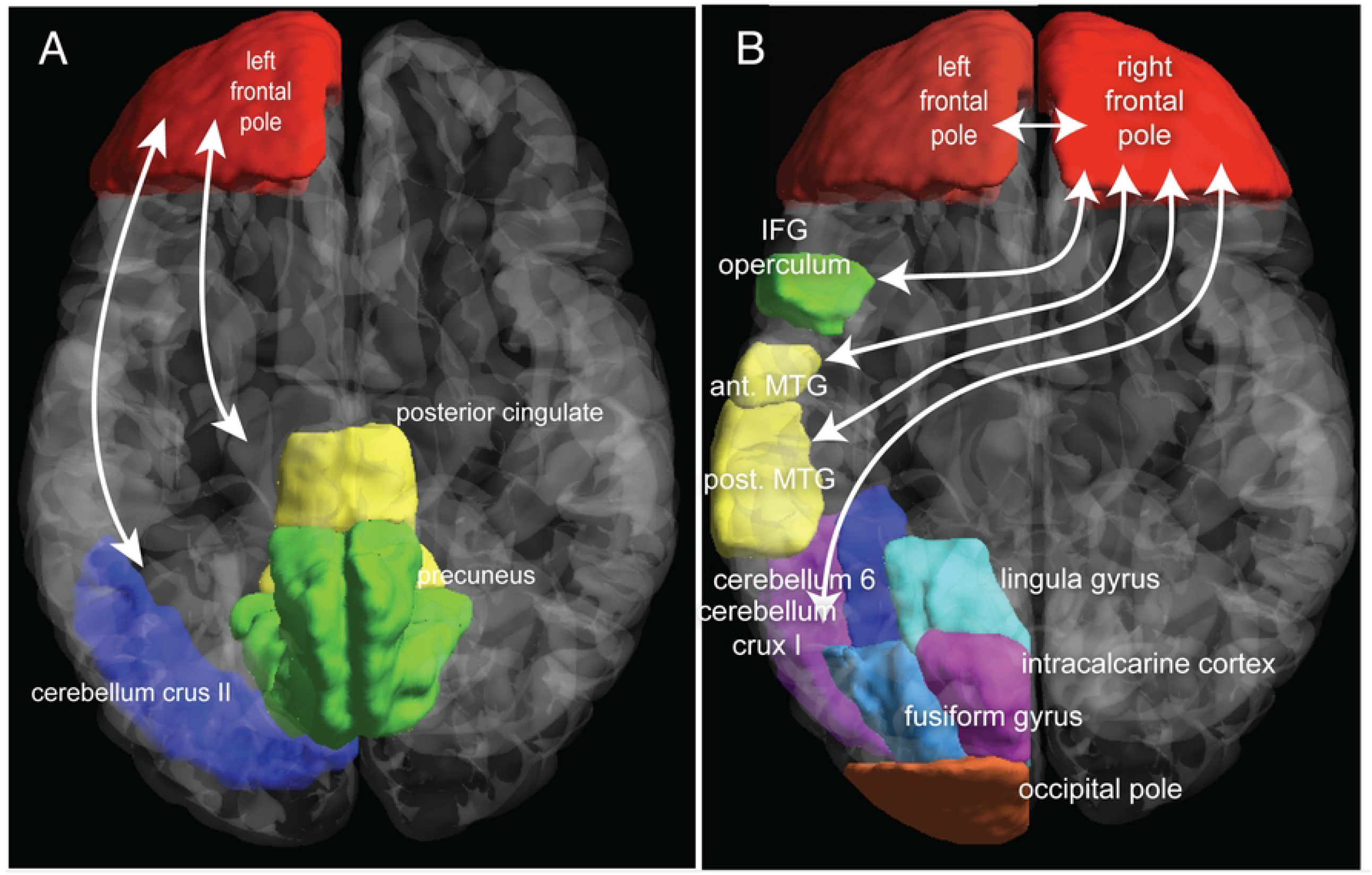
Illustration of the location of regions with increased functional connectivity to frontal poles following use of bright light therapy. **A**. Percent bright light therapy use and enhanced functional connectivity between left frontal pole and two clusters: one containing the precuneus and posterior cingulate cortex and the other the cerebellum crus2. **B**. Percent bright light therapy use and enhanced functional connectivity between right frontal pole and five clusters. The largest included cerebellum crus1, cerebellum 6, occipital pole, lingula gyrus, fusiform gyrus and intracalcarine cortex. The other clusters contained the inferior frontal gyrus (IFG) operculum, anterior and posterior middle temporal gyri (MTG) and right and left frontal poles.

### Structural MRI

Mixed effects models were used to ascertain if there was a linear or quadratic effect of device use on within subject right and left dentate gyrus volume, based on the 11 participants with pre/post structural scans. The best fitting model indicated that there was a significant quadratic effect of percent BLT use on the right dentate gyrus (F_1,7_ = 6.13, p = .043), which showed a trend level interaction with age (F_1,7_ = 3.57, p = .100). There was no significant effect of device use on left dentate gyrus volume. This was a prespecified hypothesis and no other regions were analyzed.

### Adverse Events

None of the participants reported any adverse events from use of the BLT device of blue-light blocking glasses.

## Discussion

Degree of BLT device use in this sample of healthy albeit often drowsy adolescents correlated positively with advances in their rise time, particularly on non-school days where there was more flexibility and resulted in more consistent rise times throughout the week. There was also a trend level association between degree of device use and improvement in sleep efficiency and reduction in self-reported sleepiness. As predicted, participants who used the device more frequently showed improvement in attention performance with enhanced accuracy due to a substantial reduction in errors of omission on the cognitive control attention task, reduced response variability on the SAST and improved mathematical performance.

EEG studies showed that degree of device use correlated significantly with extent of beta1, beta2 and gamma spectral power in right and left frontal poles during eyes-open testing and with beta spectral power in Fz during eyes-closed testing, consistent with enhanced frontal cortical activity and increased wakefulness. These EEG findings were supported by intrinsic functional connectivity fMRI analyses focusing on the frontal poles. There were enhanced interconnections between frontal and occipital regions and frontal – cerebellar regions. These connections make sense as the subjects were engaged in a Go / No-Go cognitive control task in which they had to visually attend to stimuli that appeared in random screen positions and based on shape either rapidly push a button or inhibit a prepotent response. Hence, the task required communication between frontal and occipital regions for discriminating targets from non-targets and likely recruited cerebellum both for its role in cognition [71, 72] and in motor response. In addition, there was increase in connectivity between left frontal pole and the posterior cingulate cortex and precuneus. These are key components of the default mode, which is active when individuals are at rest, daydreaming or ruminating. Coordinated deactivation of this system is critical for sustained attention [73], and this capacity is impaired by sleep deprivation [74]. Connectivity was also increased between the right frontal pole and medial temporal gyrus, which aids in the processing of quickly presented complex visual phenomenon [75] and the inferior frontal gyrus operculum, which is part of Broca’s area and is also involved in inhibitory control [76]. The frontal poles are the brain regions that have expanded to the greatest degree in humans. They play an important role in emotional regulation and in cognitive abilities, such as action selection [77] and have been reported to be involved in monitoring of outcomes in Go/No-Go tasks [78-80].

There were no significant associations between degree of BLT use and theta activity, which we predicted would be reduced by BLT. We suspect that participants in this study were, on average, drowsy to only a modest degree which was reflected at baseline in a reduction in frontal beta spectral activity but not in a substantial increase in theta activity.

Interestingly, there was a significant association between degree of BLT device use and size of the dentate gyrus. This was not an incidental finding, but a planned comparison based on preclinical studies [64]. We collected both T1 and T2 morphometry to provide FreeSurfer with optimal data for segmenting this region and we limited our statistical comparison to just this structure. In this case we found a significant curvilinear correlation between degree of use and change in right-sided volume. Although it is not possible to assess degree of neurogenesis from a structural scan, enhanced neurogenesis provides a plausible explanation for an increase in volume as the dentate gyrus is recognized as the primary region of the human brain with extensive postnatal neurogenesis, though the extent to which this persists beyond childhood is still debated [81]. The dentate gyrus plays a critical role in explicit memory formation and retrieval and may specifically aid in learning new material in familiar setting and discriminating old from new memories [82].

In short, this study provides evidence to support the proposition that morning use of bright LED light can enhance morning wakefulness in school children who reported feeling drowsy during early morning classes but were not specifically suffering from delayed sleep phase syndrome. We were careful to perform all assessments during morning hours (7 – 9 AM) to coincide with times when they typically felt drowsy. This potential effect of morning light is consistent with studies that that report an ‘energizing’ or alertness-enhancing effect of bright morning light [31, 83-85] but it is does not appear that this association can be accounted for by an earlier sleep onset or increased total sleep time.

Delaying school start time by about an hour is another strategy for enhancing alertness and academic performance in adolescents, and this approach has discernible benefits on sleep duration and quality [86-89]. Hence, this strategy may be even more beneficial than bright light treatment, but it requires the cooperation of the school district and may be impractical in many communities. A direct comparison between these strategies would be enlightening.

The key limitations of this study were modest sample size, particularly for the MRI component, and the open unblind design. To deal with the unblind design we surreptitiously collected data on degree of BLT device use to test the hypothesis that there would be a significant dose-response relationship between degree of use and outcome. Dose-response studies provide some of the strongest evidence for a causal relationship (i.e., Hill criteria for causality [90]). However, that is true when individuals are randomly assigned to experience different doses. Voluntary selection of dose is less powerful as we do not know for certain whether subjects who used it more benefitted more, or if subjects who experienced more benefit used it more reliably. The fact that there were alterations in objective measures (Quotient test, EEG, MRI, fMRI, actigraphy) argues for a therapeutic effect as objective measures are less susceptible to placebo response than subjective measures such as rating scales [91-94]. Nevertheless, randomized control trials are necessary to establish efficacy.

Ten or eleven repeated MRI scans is a small sample for gauging effects of treatment and should be regarded as preliminary. It is not however unusual. According to Szucs & Ioannidis [95] 96% of highly cited experimental fMRI studies had a single group of participants and these studies had a median sample size of 12 participants. With 11 participants a repeated measures study would only have sufficient power (1-β = .8) at α = .05 to detect a large effect size difference (f = .47) assuming a correlation of .5 between pre and post measures [96]. Pre and post BLT volume measures for the dentate gyrus correlated .86, so this analysis had sufficient power to detect a medium effect size (f = .25) difference. We sought to minimize false positives by restricting the volumetric analysis to the dentate gyrus and the functional connectivity analyses to the frontal poles. Nevertheless, these findings should be regarded cautiously and require replication with larger samples.

A third limitation is that actigraphs cannot estimate sleep stages [97]. Hence, we do not know if use of the device increased proportion of time spent in REM or slow wave sleep. This may be key to understanding the possible influence of bright light on cognition as well as dentate gyrus volume. A statistically significant effect of use may have emerged because sleep deprivation (particularly REM) suppresses neurogenesis in rats [98] through elevations in glucocorticoid levels [99]. Further, reduced sleep efficiency is associated with smaller dentate gyrus volumes in humans [100]. Restoring sleep after a period of deprivation produces a rebound overshoot in dentate gyrus neurogenesis [99]. Hence, the possible effect of BLT on dentate gyrus volume may have been mediated by unmeasured effects on sleep architecture.

A fourth limitation is that we restricted the cognitive measures to those that were highly dependent on attention, vigilance and mathematical performance. It would also have been useful to have included memory tests, particularly those thought to be sensitive or dependent on hippocampal function.

A final limitation is that no data were collected on use of the blue light blocking glasses, so we do not know how much influence their use might have had. A recent study using experience sampling found no association between bedtime use of social media (with the accompanying light exposure) and actigraph-assessed sleep measures [101], suggesting that the blue-ray blocking glasses may not be of primary importance.

Overall, this study provided initial evidence that voluntary degree of use of an LED BLT device and blue-ray blocking glasses at night enhanced electrophysiological correlates of alertness and measures of vigilance in a dose-dependent manner in a select group of high school students who felt sleepy during morning classes. The sample was predominantly white and enrolled in a private high school. How well these findings might generalize to a more diverse population is unclear. There is some evidence that black subjects have larger phase advances and smaller phase delays in response to bright light pulses than white participants [102]. While randomized control data are needed, particularly in larger and more representative samples, the use of BLT may make sense as there appeared to be significant benefits in this open study and there is little downside to the use of a light device that is no brighter than what one would experience looking out the window on a spring morning.

## Data Availability

All relevant data except the MRI scans are within the manuscript and its Supporting Information files. MRI scans have been uploaded to OpenNeuro doi:10.18112/openneuro.ds004219.v1.0.0

doi:10.18112/openneuro.ds004219.v1.0.0

## Acknowledgment

We thank Rosalind Gendreau, R.N., M.S.N., who was the Director of Health Services at the Walnut Hill School. She provided invaluable assistance in organizing the study and enabling it to take place at Walnut Hill.

